# Multitask Artificial Intelligence–Based Electrocardiogram Tool for Preoperative Cardiac Testing in Noncardiac Surgery: Retrospective Cohort Study of Health Care Utilization and Costs

**DOI:** 10.64898/2025.12.21.25342775

**Authors:** Hong-Mi Choi, Yerin Kim, Joonghee Kim, Jiesuck Park, In-Chang Hwang, Yun Young Choi, Ji Hyun Lee, Yeonyee E. Yoon, Il-Young Oh, Goo-Yeong Cho, In-Ae Song, Youngjin Cho

## Abstract

**Background:** Preoperative cardiovascular (CV) risk stratification is essential in non-cardiac surgery, but conventional testing is frequently overused, increasing costs without improving outcomes. Artificial intelligence (AI)-enabled electrocardiography (ECG) may enhance perioperative risk assessment by identifying patients at very low risk for adverse events.

**Objective:** This study aimed to evaluate whether AI-ECG-based risk stratification could help maintain safety and decrease potentially avoidable preoperative CV testing, while reducing the associated costs, in patients undergoing non-cardiac surgery.

**Methods:** We retrospectively analyzed 41,218 patients (46,135 ECG–surgery pairs) undergoing elective non-cardiac surgery at Seoul National University Bundang Hospital (2020–2021). An AI-ECG algorithm generated eight probability scores for cardiac conditions, classifying patients as low- or high-risk. Based on the performance and results of preoperative cardiovascular testing (transthoracic echocardiography, coronary computed tomography angiography, single-photon emission computed tomography, or coronary angiography), patients were classified as no test, negative test, or positive test. The primary endpoint was a 30-day composite of all-cause mortality, unplanned percutaneous coronary intervention, or prolonged mechanical ventilation (≥3 days).

**Results:** AI-ECG classified 92.4% of patients as low-risk, with an event rate of 0.62% versus 6.04% in high-risk patients. Preoperative CV testing was performed in 11.8% of cases, with only 16.3% yielding positive findings. In AI-ECG low-risk patients, event rates were uniformly low (0.6–2.7%) regardless of testing, whereas in high-risk patients, rates were consistently high (5.3–6.2%), suggesting no additional prognostic value of conventional testing. An AI-ECG guided approach could reduce potentially avoidable preoperative testing by 35.7% while preserving safety. Integrating AI-ECG with established risk tools may better delineate patients who truly require preoperative CV testing while conserving medical resources.

**Conclusion:** AI-enabled ECG reliably identified surgical candidates at low risk for postoperative complications, for whom additional CV testing may be potentially avoidable. Integrating AI-ECG with conventional risk tools may optimize resource use and minimize redundant testing without compromising outcomes. Prospective studies are needed to confirm clinical and economic benefits.

**Trial Registration:** N/A

## Introduction

More than 200 million non-cardiac surgeries are conducted worldwide each year,[1] with numbers steadily increasing.[2] Preoperative cardiovascular (CV) testing imposes a major healthcare burden, with estimated costs of $18 billion annually in the United States.[3] The substantially higher morbidity and mortality risks of postoperative CV complications underscore the importance of accurate preoperative risk stratification.[4] Although existing risk-estimation tools provide general estimates of perioperative CV risk,[5, 6] they do not specify whether additional testing is warranted and do not eliminate the risk of adverse outcomes. Despite current guidelines outlining which patients may benefit from advanced CV testing,[5, 6] in clinical practice, the demand for such testing often exceed the recommended indications.[7] As a result, preoperative CV tests are frequently overused and thereby hindering efficient healthcare resource utilization.

Electrocardiography (ECG) is a well-established low-cost preoperative assessment tool, but has limited utility in non-cardiac surgery.[3, 8, 9] Not surprisingly, the quality of ECG-derived information has been highly dependent on the physician’s expertise and the patient characteristics.[10, 11] Thus, recent guidelines have recommended ECG use in only selected patients with suspected CV diseases or risk factors who are undergoing intermediate- or high-risk non-cardiac surgery.[5, 6] However, recent advances in artificial intelligence (AI) have expanded the diagnostic and prognostic applications of ECG, enabling detection of coronary artery disease, valvular heart disease, and myocardial dysfunction.[12, 13] With its low-cost, broad accessibility, AI-enhanced ECG is a promising tool for preoperative risk stratification.

Building on a pre-existing AI-ECG model that incorporated eight quantitative scores for ECG (QCGs) reflecting diverse cardiac conditions,[14–19] we aimed to evaluate whether AI-ECG-based risk stratification could help decrease potentially avoidable preoperative CV testing, while reducing the associated costs and maintaining safety in patients undergoing non-cardiac surgery. We hypothesized that the integration of AI-ECG into the workflow of preoperative risk assessment could help identify a subgroup of patients with an extremely low risk of postoperative adverse outcomes and thereby warrant no additional testing.

## Materials and methods

### Study population

This single-center, retrospective cohort study included patients who underwent elective non-cardiac surgery at Seoul National University Bundang Hospital between 2020 and 2021 and underwent an ECG within the 30 days preceding the surgery. The exclusion criteria included: 1) no preoperative ECG within 30 days preceding the surgery; 2) elective cardiac surgery or extracorporeal membrane oxygenation-related surgery; 3) the use of simple pain-control procedures; 4) surgeries performed under local anesthesia; 5) surgery for a deceased organ donor; 6) rigid bronchoscopy; 7) dental surgery; and 8) multiple surgeries during the index admission. A total of 46,135 ECG–surgery pairs from 41,218 patients were analyzed (**Figure S1** in Multimedia Appendix).

### Data collection

All ECGs were recorded under standardized settings (25 mm/s speed and 10 mm/mV gain) and processed to extract the QCG scores from the JPEG file. The preoperative ECG that was recorded closest to the surgery was paired with one non-cardiac surgery. Clinical data, including surgery-related records, test results, and outcomes, were obtained from electronic health records (EHRs). This observational study conformed to the STROBE reporting guideline.[20] The study protocol was approved by the Institutional Review Board of Seoul National University Bundang Hospital (IRB No. B-2409-927-106), which waived the requirement for written informed consent owing to the retrospective nature of the study. All the clinical investigations were conducted in accordance with the principles of the Declaration of Helsinki.

### AI algorithm and risk-stratification strategy for AI-ECG

The QCG system is a deep-learning-based AI analyzer that uses 12-lead ECG images (in JPEG, PNG, or PDF formats) as input data and yields the probability score of specific downstream tasks as numerical values (range 0–100). Following pretraining using self-supervised learning schemes, these models were fine-tuned using multi-task learning schemes with a modified convolutional neural network using residual connections, squeeze excitation modules, and a nonlocal block. The input data comprised 47,194 annotated ECG images from Seoul National University Bundang Hospital that were recorded between 2017 and 2019. The tasks included rhythm classification and the construction of ten QCG scores related to various medical emergencies: 1) critical illness (critical score); 2) acute coronary syndrome (ACS); 3) ST-segment elevation myocardial infarction (STEMI); 4) myocardial injury; 5) pulmonary edema; 6) pericardial effusion; 7) left ventricular dysfunction; 8) right ventricular dysfunction; 9) pulmonary hypertension; and 10) hyperkalemia. ECG buddy—an application that provides these 10 QCG scores—was approved by the Korean Ministry of Food and Drug Safety (January 2024). Detailed explanations of the AI algorithms and validation studies have been previously published.[15–18] The detailed explanation of each QCG score is provided in **Table S1** in Multimedia Appendix.

Among the ten probability scores, we selected eight QCG scores that are directly related to the cardiac conditions of interest, except for the critical and hyperkalemia scores. All scores had their own optimal thresholds to maximize diagnostic performance, which were determined at the model-development stage. The participants were assigned to the AI-ECG low-risk group when all 8 QCG scores are lower than their individual optimal thresholds; otherwise, they were assigned to the AI-ECG high-risk group.

### Preoperative cardiovascular tests

Preoperative cardiovascular tests included echocardiography, coronary computed tomography angiography (CCTA), single-photon emission computed tomography (SPECT) performed within 90 days before surgery, and coronary angiography (CAG) performed within 180 days before surgery. The decision for further preoperative CV testing and the choice of diagnostic modality was made by the attending surgeon or consulting physician while considering each patient’s functional capacity, individual risk, and surgical risk. Although routine clinical practice already includes these tests irrespective of the surgery, all CV tests performed within the designated time window were considered preoperative CV testing.

The positive findings of preoperative CV tests were selected based on widely accepted actionability, whereby positive results suggested the need for further preoperative evaluation. For echocardiography, positive findings were defined as the presence of any of the following: moderate or severe valvular heart disease (aortic stenosis or regurgitation, mitral stenosis or regurgitation, or tricuspid regurgitation); left ventricular ejection fraction ≤50%; or regional wall motion abnormality. For CCTA and CAG, ≥50% stenosis in any of the three major coronary arteries was considered a positive result; angiographic stenosis grades were used for CAG. For SPECT, a ≥10% reversible or irreversible perfusion defect was defined as a positive result. The costs of CCTA, CAG, and SPECT were obtained from a published study conducted in the Republic of Korea in 2018 that reported values in U.S. dollars.[21] For echocardiography, the cost was based on the median value of publicly posted prices in Seoul, Republic of Korea, and was converted from KRW to U.S. dollars using an approximate 2021 exchange rate.[22]

Based on the preoperative CV testing status, all the surgery–ECG pairs were categorized into three groups: No test, patients without any preoperative CV test; Negative test, patients who underwent testing without any positive findings; and Positive test, patients with at least one positive finding. In this context, a negative test indicates the absence of predefined positive findings in a specific examination rather than the confirmation of overall clinical normality.

### Study group definition

To compare the adequacy of the preoperative CV testing and AI-enabled ECG, the study population was further divided into six groups: (1) Group 1, low-risk AI-ECG and No test; (2) Group 2, low-risk AI-ECG and Negative test; (3) Group 3, low-risk AI-ECG and Positive test; (4) Group 4, high-risk AI-ECG and No test; (5) Group 5, high-risk AI-ECG and Negative test; and (6) Group 6, high-risk AI-ECG and Positive test (**Figure 1**).

**Figure 1.**
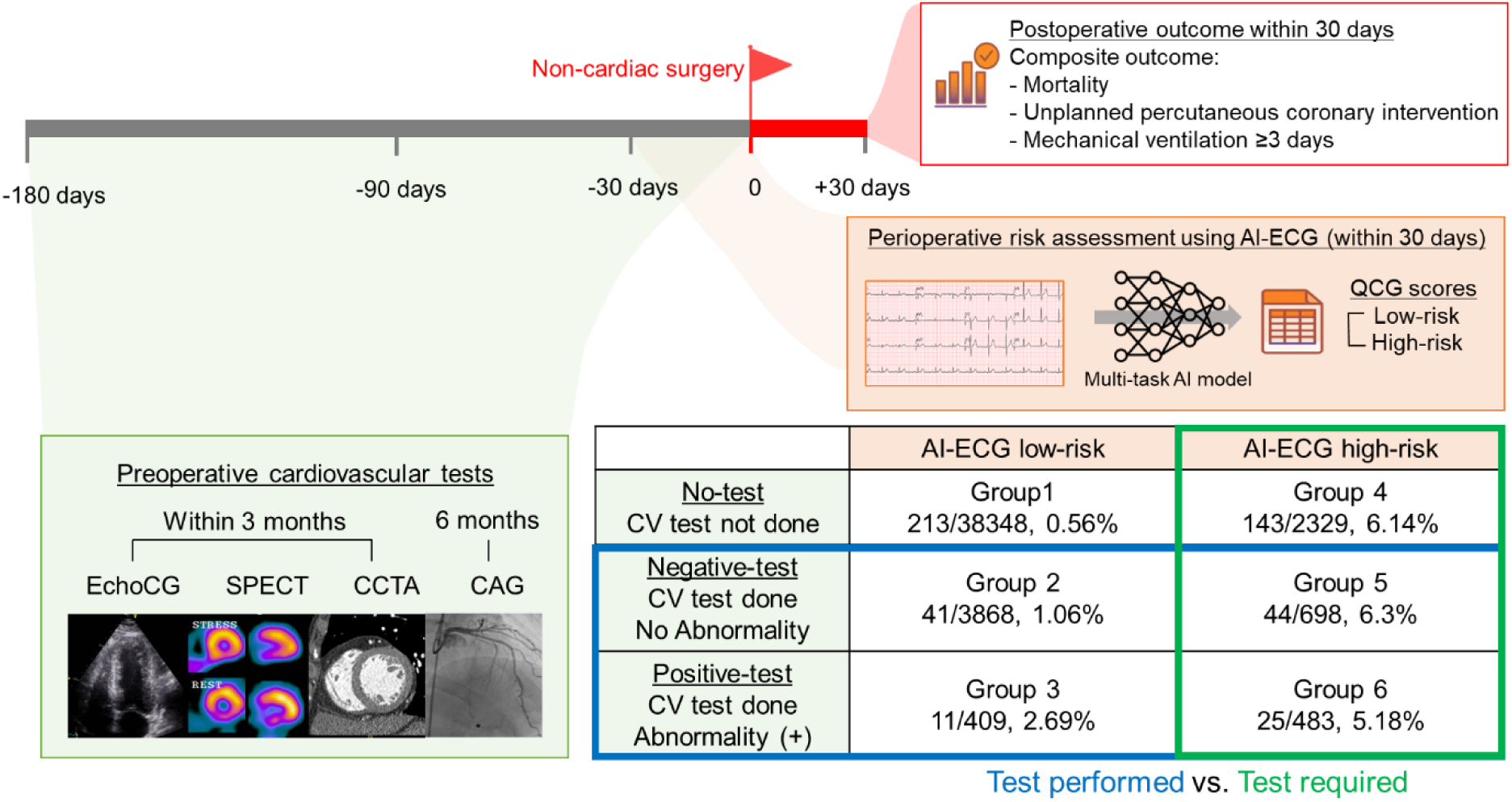
Study design. Among the preoperative cardiovascular (CV) tests, we included echocardiography (EchoCG), single-photon emission computed tomography (SPECT), and coronary computed tomography angiography (CCTA) that were performed within 90 days, and coronary angiography (CAG) performed within 180 days before the non-cardiac surgery. Among the electrocardiography (ECG) scans performed within 30 days preoperatively, the ECG obtained nearest to the surgery was selected to estimate the QCG scores using an artificial intelligence (AI)-enabled ECG. Next, the preoperative risk was classified using eight QCG scores related to cardiac conditions. Six groups were created based on the preoperative CV test and AI-ECG risk classification. The blue square denotes the surgeries in which CV tests were performed (groups 2, 3, 5, and 6; blue square), and the green square implies the surgeries in which testing would have been required (groups 4, 5, and 6; green square) assuming that the decision to perform preoperative CV testing strictly followed the AI-ECG risk stratification (i.e., tests performed only in high-risk patients).

### Postoperative outcome

The primary measure that was used to assess the predictive performance of preoperative CV testing and AI-enabled ECG was a composite postoperative outcome occurring within 30 days after the index surgery that included: (1) all-cause mortality; (2) unplanned percutaneous coronary intervention; and (3) mechanical ventilation for ≥3 days. Mortality was restricted to deaths during hospitalization, including deaths within 30 days of readmission, whereas deaths that occurred outside the hospital were not included in the analysis. The overall study scheme is illustrated in **Figure 1**.

### Statistical analysis

Continuous variables were summarized as means with standard deviations or medians with interquartile ranges whereas categorical variables were summarized as frequencies with percentages. Between-group comparisons were performed using the independent two-sample *t*-test or the Mann–Whitney *U* test, for continuous variables, and the chi-square or Fisher’s exact test, for categorical variables, as appropriate.

For intergroup comparison, the incidence of the composite outcome was compared between the AI-ECG high- and low-risk groups within the RCRI and ESC surgical risk strata, and the incidence rates with 95% confidence intervals (CIs), relative risks (RRs), and risk differences (RDs) were derived from contingency 2×2 tables comparing AI-ECG risk groups and the presence or absence of the composite outcome. Within each AI-ECG stratum, the outcome incidence across the three conventional testing groups (No test, Negative test, and Positive test) was compared using the chi-square or Fisher’s exact tests; pairwise Fisher’s exact tests and Bonferroni correction applied when appropriate. For comparisons across multiple strata, overall differences were evaluated using one-way ANOVA or the Kruskal– Wallis test, for continuous variables, and the chi-square or Fisher’s exact test, for categorical variables.

Statistical significance was defined using a two-sided *p*-value <0.05. All analyses were performed using R, version 4.4.1 (https://www.R-project.org).

## Results

### Baseline characteristics

Baseline clinical and surgery-specific characteristics of the study population are summarized in **Table 1**. The mean age was 56.6 (SD, 16.1) years, and 44.6% of the patients were male. Emergency procedures comprised 6.1% of all surgeries, and 72.4% were performed under general anesthesia. According to the European Society of Cardiology (ESC) surgical risk categorization and the Revised Cardiac Risk Index (RCRI) stratification, high-risk surgery was identified in 10.7% and 4.2% of patients, respectively.

**Table 1.** Baseline characteristics and comparison between AI-ECG risk groups.

|  | Overall<br>(N=46,135) | AI-ECG low<br>risk<br>(N=42,625) | AI-ECG<br>high-risk<br>(N=3,510) | p |
| --- | --- | --- | --- | --- |
| *Demographics and comorbidities* |  |  |  |  |
| Age (mean (SD)) | 56.56 (16.14) | 55.66 (15.98) | 67.37 (14.00) | <0.001 |
| Male sex (%) | 20561 (44.6) | 18302 (42.9) | 2259 (64.4) | <0.001 |
| DM (%) | 5904 (12.8) | 4918 (11.5) | 986 (28.1) | <0.001 |
| DM treated with insulin (%) | 632 (1.4) | 445 (1.0) | 187 (5.3) | <0.001 |
| Hypertension (%) | 10759 (23.3) | 9311 (21.8) | 1448 (41.3) | <0.001 |
| Previous heart failure (%) | 410 (0.9) | 141 (0.3) | 269 (7.7) | <0.001 |
| Previous ischemic heart disease (%) | 1772 (3.8) | 1141 (2.7) | 631 (18.0) | <0.001 |
| Previous cerebrovascular accident (%) | 1525 (3.3) | 1216 (2.9) | 309 (8.8) | <0.001 |
| Smoking (%) |  |  |  |  |
| Nonsmoker | 28436 (61.6) | 26356 (61.8) | 2080 (59.3) | <0.001 |
| Smoker | 9904 (21.5) | 8782 (20.6) | 1122 (32.0) |  |
| Unknown | 7795 (16.9) | 7487 (17.6) | 308 (8.8) |  |
| Creatinine (median [IQR]) | 0.74 [0.62, 0.91] | 0.73 [0.61, 0.89] | 0.90 [0.70, 1.26] | <0.001 |
| Creatinine $\geq 2.0$ mg/dl (%) | 1414 (3.1) | 857 (2.0) | 557 (15.9) | <0.001 |
| *Surgery-related* |  |  |  |  |
| Emergency surgery (%) | 2806 (6.1) | 2340 (5.5) | 466 (13.3) | <0.001 |
| Anesthesia (%) |  |  |  |  |
| General anesthesia | 33386 (72.4) | 30960 (72.6) | 2426 (69.1) | <0.001 |
| Spinal/Epidural anesthesia | 4272 (9.3) | 3965 (9.3) | 307 (8.7) |  |
| Monitored anesthesia care | 8477 (18.4) | 7700 (18.1) | 777 (22.1) |  |
| ESC surgical risk category (%) |  |  |  |  |
| Low | 24825 (53.8) | 23296 (54.7) | 1529 (43.6) | <0.001 |
| Intermediate | 16392 (35.5) | 15166 (35.6) | 1226 (34.9) |  |
| High | 4918 (10.7) | 4163 (9.8) | 755 (21.5) |  |
| RCRI (%) |  |  |  |  |
| 0 | 28738 (62.3) | 27427 (64.3) | 1311 (37.4) | <0.001 |
| 1 | 15468 (33.5) | 14019 (32.9) | 1449 (41.3) |  |
| 2 | 1562 (3.4) | 1023 (2.4) | 539 (15.4) |  |
| 3 | 308 (0.7) | 141 (0.3) | 167 (4.8) |  |
| 4 | 50 (0.1) | 15 (0.0) | 35 (1.0) |  |
| 5 | 9 (0.0) | 0 (0.0) | 9 (0.3) |  |
| ASA classification (%) |  |  |  |  |
| 1 | 13814 (29.9) | 13623 (32.0) | 191 (5.4) | <0.001 |
| 2 | 24130 (52.3) | 22955 (53.9) | 1175 (33.5) |  |
| 3 | 7332 (15.9) | 5612 (13.2) | 1720 (49.0) |  |
| 4 | 765 (1.7) | 382 (0.9) | 383 (10.9) |  |
| 5 | 94 (0.2) | 53 (0.1) | 41 (1.2) |  |
| *QCG scores (median [IQR])* |  |  |  |  |
| ACS | 1.37 [0.30, | 1.20 [0.27, | 12.29 [2.82, | <0.001 |
|  | 4.26] | 3.62] | 25.80] |  |
| STEMI | 0.01 [0.00, 0.07] | 0.01 [0.00, 0.05] | 0.40 [0.06, 1.89] | <0.001 |
| Myocardial injury | 1.71 [0.50, 4.58] | 1.49 [0.44, 3.80] | 16.71 [6.42, 26.07] | <0.001 |
| Pulmonary edema | 0.63 [0.20, 2.18] | 0.55 [0.18, 1.72] | 15.23 [2.76, 26.96] | <0.001 |
| Pericardial effusion | 0.03 [0.00, 0.23] | 0.02 [0.00, 0.16] | 1.34 [0.10, 7.41] | <0.001 |
| Left ventricular dysfunction | 0.08 [0.02, 0.34] | 0.07 [0.02, 0.24] | 4.10 [0.70, 19.05] | <0.001 |
| Right ventricular dysfunction | 0.13 [0.04, 0.57] | 0.11 [0.03, 0.41] | 4.99 [0.74, 17.64] | <0.001 |
| Pulmonary hypertension | 0.15 [0.03, 0.76] | 0.12 [0.03, 0.56] | 5.60 [0.62, 17.62] | <0.001 |
| *Preoperative cardiovascular test* |  |  |  |  |
| Any preoperative CV test performed (%) | 5458 (11.8) | 4277 (10.0) | 1181 (33.6) | <0.001 |
| Positive any preoperative CV test (%) | 892 (16.3) | 409 (9.6) | 483 (40.9) | <0.001 |
| Echocardiography performed (%) | 4808 (10.4) | 3846 (9.0) | 962 (27.4) | <0.001 |
| Positive echocardiography results (%) | 574 (11.9) | 246 (6.4) | 328 (34.1) | <0.001 |
| Moderate or severe valvular heart disease (%) | 235 (4.9) | 114 (3.0) | 121 (12.7) | <0.001 |
| Regional wall motion abnormality (%) | 374 (7.8) | 133 (3.5) | 241 (25.3) | <0.001 |
| LVEF < 50% (%) | 225 (4.7) | 55 (1.4) | 170 (17.8) | <0.001 |
| LV ejection fraction (%) (median [IQR]) | 62.66 [58.90, 66.34] | 63.16 [59.74, 66.67] | 60.00 [54.05, 64.78] | <0.001 |
| Coronary angiography performed (%) | 470 (1.0) | 209 (0.5) | 261 (7.4) | <0.001 |
| Positive coronary angiography results (%) | 273 (58.1) | 102 (48.8) | 171 (65.5) | <0.001 |
| Coronary CT angiography performed (%) | 579 (1.3) | 414 (1.0) | 165 (4.7) | <0.001 |
| Positive coronary CT angiography results (%) | 165 (29.0) | 102 (24.9) | 63 (39.4) | 0.001 |
| SPECT performed (%) | 245 (0.5) | 153 (0.4) | 92 (2.6) | <0.001 |
| Positive SPECT results (%) | 7 (2.9) | 3 (2.0) | 4 (4.3) | 0.49 |

Overall, 3,510 patients (7.6%) had at least one QCG value that exceeded the predefined cutoff and were thus classified as high-risk by AI-ECG. Compared to the low-risk group, high-risk patients were older, included more males, had a greater burden of comorbidities, underwent a higher proportion of emergency procedures, and were more frequently categorized as undergoing high-risk surgeries, according to both the ESC and RCRI (**Table 1**) risk categorizations.

The distribution of the QCG scores across the six study groups is shown in **Figure S2** in Multimedia Appendix, with significantly higher scores observed in the AI-ECG-stratified high-risk group. Across the three strata of preoperative CV test results, patients in the Positive-test group were the oldest, had the highest comorbidity burden, underwent high-risk surgery, and had the highest QCG scores. Patients in the Negative-test group showed intermediate characteristics, whereas those in the No-test group were the youngest, with the fewest comorbidities and the lowest QCG scores (**Table S2** in Multimedia Appendix).

### Postoperative outcomes

In total, 477 patients (1.0%) experienced the postoperative composite outcome, with prolonged mechanical ventilation (≥3 days) being the most frequent component. Overall, the event rates were 6.04% and 0.62% in the high-risk and low-risk AI-ECG groups, respectively. When stratified by surgical urgency, composite outcomes occurred in 9.7% and 0.5% of emergency and elective surgeries, respectively. Among the patients undergoing emergency surgery, the risks of the composite endpoint were 6.5% and 25.8% in the AI-ECG-stratified low-risk and high-risk groups, respectively.

The incidence of the composite outcome was consistently higher in AI-ECG-graded high-risk patients across all ESC surgical risk and RCRI strata. In the ESC surgical risk- and RCRI-categorized low-risk procedures, the RRs of the composite outcome occurrence were higher in the high-risk AI-ECG group than low-risk AI-ECG group (RR [95% CI] 28.57 [15.61–52.29] and 9.21 [7.49–11.31], respectively). The procedures classified as high-risk by the established risk stratification tools showed larger RDs between the low-risk and high-risk AI-ECG groups (**Table 2 and Figure S3** in Multimedia Appendix).

**Table 2.** The incidence, relative risk, and relative difference of the composite outcome across ESC surgical risk category and RCRI strata by AI-ECG risk stratification.

|  | Risk group | Incidence of composite outcomes |  |  |  |
| --- | --- | --- | --- | --- | --- |
|  |  | AI-ECG-stratified low-risk (%) | AI-ECG-stratified high-risk (%) | RR (95% CI) | RD %p (95% CI) |
| Surgical Emergency | Elective | 0.28 (0.23–0.34) | 3.02 (2.44–3.69) | 10.68 (8.14–14.02) | 2.74 (2.13–3.35) |
|  | Emergency | 6.45 (5.49–7.53) | 25.75 (21.84–29.97) | 3.99 (3.21–4.96) | 19.30 (15.21–23.39) |
| ESC surgical risk | Low | 0.07 (0.04–0.11) | 1.96 (1.33–2.79) | 28.57 (15.61–52.29) | 1.89 (1.20–2.59) |
|  | Intermediate | 1.09 (0.94–1.27) | 6.44 (5.13–7.97) | 5.89 (4.53–7.65) | 5.35 (3.96–6.73) |
|  | High | 1.99 (1.59–2.47) | 13.64 (11.27–16.30) | 6.84 (5.18–9.04) | 11.65 (9.16–14.13) |
| RCRI | 0-1 | 0.55 (0.49 to 0.63) | 5.11 (4.32 to 6.00) | 9.21 (7.49–11.31) | 4.55 (3.73–5.38) |
|  | 2+ | 2.97 (2.08–4.10) | 9.47 (7.47–11.79) | 3.19 (2.15–4.73) | 6.50 (4.19–8.81) |
| Total |  | 0.62 (0.55–0.70) | 6.04 (5.27–6.88) | 9.72 (8.14–11.60) | 5.42 (4.63–6.21) |

CI, confidence interval; ESC, European Society of Cardiology; RCRI, Revised Cardiac Risk Index; RD, relative difference; RR, relative risk.

Comparisons of predefined outcomes across six study groups, defined by AI-ECG and preoperative CV test results, are summarized in **Figure 2**. The composite outcomes differed significantly between the low-risk and high-risk groups on AI-ECG (*p*<0.001 for each comparison within the No-test, Negative-test, and Positive-test groups). Low-risk AI-ECG group showed consistently low composite outcome rates compared to high-risk AI-ECG group. Among the low-risk AI-ECG patients, those who underwent conventional testing had progressively higher event rates (0.6%, 1.1%, and 2.7% in the No-test, Negative-test, and Positive-test groups, respectively; *p*<0.001). In contrast, among high-risk patients, the composite outcome incidence was uniformly high regardless of testing (6.1%, 6.2%, and 5.3%; *p*=0.683), which indicated that conventional testing did not further stratify risk after the AI-ECG had classified patients as high-risk.

**Figure 2.**
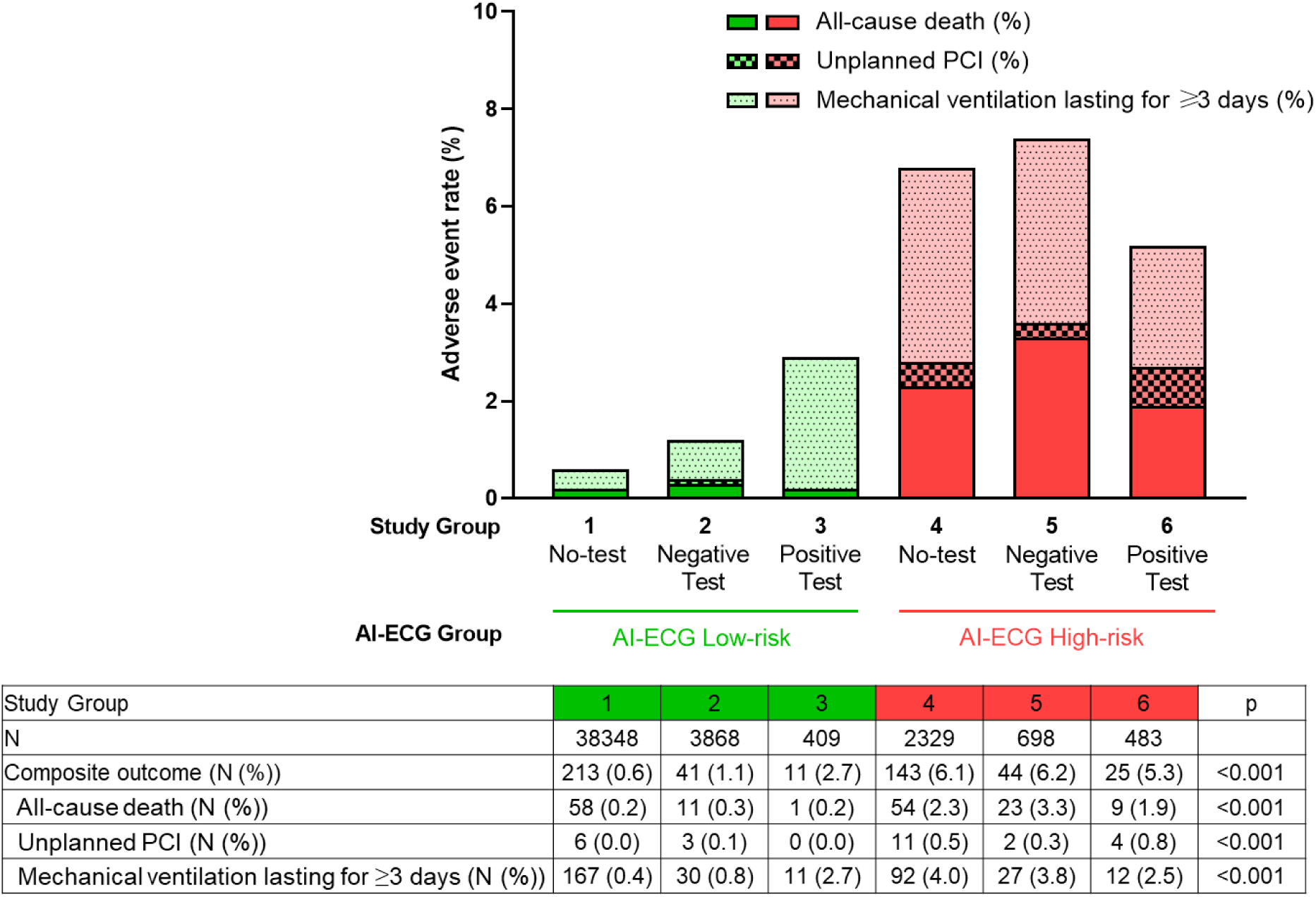
Predefined outcome rates across the six study groups assigned by the AI-ECG (Low-risk, green; High-risk, red) and the preoperative CV testing. PCI, percutaneous coronary intervention

### Preoperative CV test and reclassification using AI-ECG

Preoperative CV testing was performed in 11.8% of the cases, with only 16.3% showing positive findings. Of the patients who received preoperative cardiac testing, 88.1% underwent echocardiography, making it the most commonly used modality. Among the patients who underwent preoperative CV testing (Groups 2, 3, 5, and 6), the negative test groups (Groups 2 and 5) were less likely to undergo additional CV tests, besides echocardiography, than the positive test groups (Groups 3 and 6). The types and results of preoperative CV tests are presented in **Table S3** in Multimedia Appendix.

When stratified by preoperative CV testing, all outcomes, except for prolonged mechanical ventilation in the Positive test group, showed significant differences according to the AI-ECG risk level. As illustrated in **Figure 3**, a substantial proportion of patients in the No-test group (5.7%) were classified as high-risk by AI-ECG, whereas most patients in the Negative-test group (84.7%) were classified as low risk.

**Figure 3.**
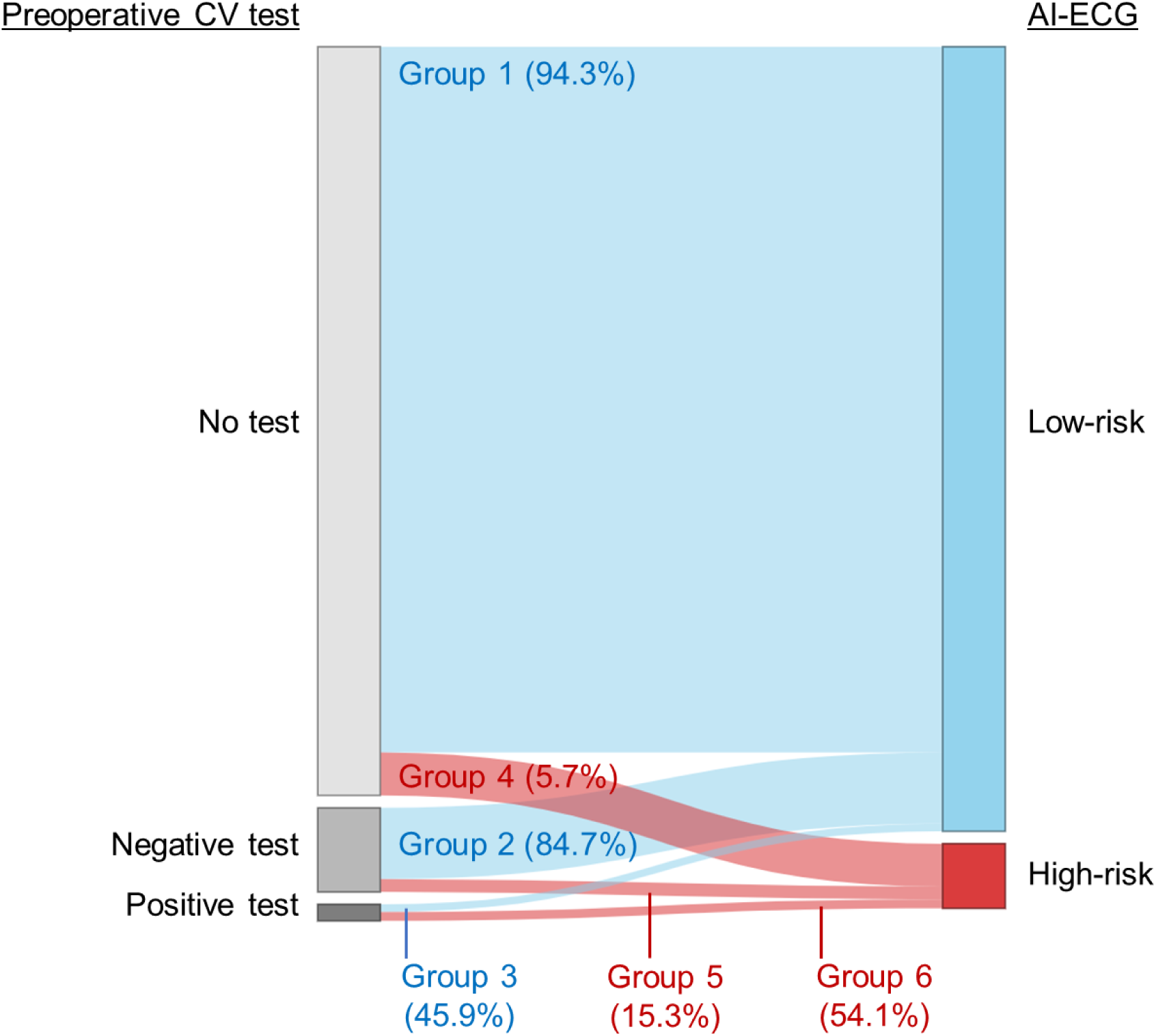
The proportion of AI-ECG risk according to the preoperative CV test. A substantial proportion of patients in the No-test group (5.7%) were classified as high risk by AI-ECG (red), whereas the majority of patients in the Negative-test group (84.7%) were classified as low risk by AI-ECG (light blue).

### Individual study groups

The characteristics of each study group are summarized in **Table 3**. Groups 1 and 6 demonstrated appropriate application of the AI-ECG–derived risk classification: Group 1 included low-risk patients who did not undergo further testing, and Group 6 included high-risk patients who underwent testing with abnormal findings. Accordingly, the subsequent evaluation focuses on Groups 2–5.

**Table 3.** Characteristics of individual study groups.

|  | Group 1<br>(N=38,348) | Group 2<br>(N=3,868) | Group 3<br>(N=409) | Group 4<br>(N=2,329) | Group 5<br>(N=698) | Group 6<br>(N=483) | p |
| --- | --- | --- | --- | --- | --- | --- | --- |
| *Demographics and comorbidities* |  |  |  |  |  |  |  |
| Age (mean (SD)) | 54.16<br>(15.69) | 68.84<br>(11.70) | 71.75<br>(10.64) | 65.25<br>(14.28) | 71.46<br>(12.87) | 71.68<br>(11.73) | <0.001 |
| Male sex (%) | 16,207<br>(42.3) | 1820<br>(47.1) | 275<br>(67.2) | 1538<br>(66.0) | 399<br>(57.2) | 322<br>(66.7) | <0.001 |
| DM (%) | 4001<br>(10.4) | 796<br>(20.6) | 121<br>(29.6) | 629<br>(27.0) | 172<br>(24.6) | 185<br>(38.3) | <0.001 |
| DM treated with insulin (%) | 315 (0.8) | 112 (2.9) | 18 (4.4) | 96 (4.1) | 47 (6.7) | 44 (9.1) | <0.001 |
| Hypertension (%) | 7557<br>(19.7) | 1553<br>(40.1) | 201<br>(49.1) | 935<br>(40.1) | 276<br>(39.5) | 237<br>(49.1) | <0.001 |
| Previous heart failure (%) | 44 (0.1) | 32 (0.8) | 65 (15.9) | 66 (2.8) | 17 (2.4) | 186<br>(38.5) | <0.001 |
| Previous ischemic heart disease (%) | 702 (1.8) | 263 (6.8) | 176<br>(43.0) | 288<br>(12.4) | 82 (11.7) | 261<br>(54.0) | <0.001 |
| Previous cerebrovascular accident (%) | 965 (2.5) | 203 (5.2) | 48 (11.7) | 199 (8.5) | 57 (8.2) | 53 (11.0) | <0.001 |
| Smoking (%) |  |  |  |  |  |  |  |
| Nonsmoker | 23402<br>(61.0) | 2735<br>(70.7) | 219<br>(53.5) | 1328<br>(57.0) | 482<br>(69.1) | 270<br>(55.9) | <0.001 |
| Smoker | 7577<br>(19.8) | 1034<br>(26.7) | 171<br>(41.8) | 722<br>(31.0) | 200<br>(28.7) | 200<br>(41.4) |  |
| Unknown | 7369<br>(19.2) | 99 (2.6) | 19 (4.6) | 279<br>(12.0) | 16 (2.3) | 13 (2.7) |  |
| Creatinine (median [IQR]) | 0.72<br>[0.61, 0.88] | 0.78<br>[0.64, 0.95] | 0.89<br>[0.74, 1.12] | 0.89<br>[0.70, 1.20] | 0.88<br>[0.68, 1.26] | 1.01<br>[0.78, 1.63] | <0.001 |
| Creatinine ≥ 2.0 mg/dl (%) | 670 (1.7) | 147 (3.8) | 40 (9.8) | 358<br>(15.4) | 96 (13.8) | 103<br>(21.3) | <0.001 |
| *Surgery-related* |  |  |  |  |  |  |  |
| Emergency surgery (%) | 2230<br>(5.8) | 88 (2.3) | 22 (5.4) | 389<br>(16.7) | 44 (6.3) | 33 (6.8) | <0.001 |
| Anesthesia (%) |  |  |  |  |  |  |  |
| General anesthesia | 27687<br>(72.2) | 2965<br>(76.7) | 308<br>(75.3) | 1576<br>(67.7) | 525<br>(75.2) | 325<br>(67.3) | <0.001 |
| Spinal/Epidural anesthesia | 3224<br>(8.4) | 698<br>(18.0) | 43 (10.5) | 163 (7.0) | 94 (13.5) | 50 (10.4) |  |
| Monitored anesthesia care | 7437<br>(19.4) | 205 (5.3) | 58 (14.2) | 590<br>(25.3) | 79 (11.3) | 108<br>(22.4) |  |
| ESC surgical risk category (%) |  |  |  |  |  |  |  |
| Low | 22098<br>(57.6) | 1080<br>(27.9) | 118<br>(28.9) | 1147<br>(49.2) | 206<br>(29.5) | 176<br>(36.4) | <0.001 |
| Intermediate | 12821<br>(33.4) | 2191<br>(56.6) | 154<br>(37.7) | 740<br>(31.8) | 322<br>(46.1) | 164<br>(34.0) |  |
| High | 3429<br>(8.9) | 597<br>(15.4) | 137<br>(33.5) | 442<br>(19.0) | 170<br>(24.4) | 143<br>(29.6) |  |
| RCRI (%) |  |  |  |  |  |  |  |
| 0 | 25503<br>(66.5) | 1855<br>(48.0) | 69 (16.9) | 1009<br>(43.3) | 256<br>(36.7) | 46 (9.5) | <0.001 |
| 1 | 12145<br>(31.7) | 1693<br>(43.8) | 181<br>(44.3) | 982<br>(42.2) | 312<br>(44.7) | 155<br>(32.1) |  |
| 2 | 624 (1.6) | 280 (7.2) | 119<br>(29.1) | 261<br>(11.2) | 104<br>(14.9) | 174<br>(36.0) |  |
| 3 | 69 (0.2) | 34 (0.9) | 38 (9.3) | 54 (2.3) | 25 (3.6) | 88 (18.2) |  |
| 4 | 7 (0.0) | 6 (0.2) | 2 (0.5) | 18 (0.8) | 1 (0.1) | 16 (3.3) |  |
| 5 | 0 (0.0) | 0 (0.0) | 0 (0.0) | 5 (0.2) | 0 (0.0) | 4 (0.8) |  |
| ASA classification (%) |  |  |  |  |  |  |  |
| 1 | 13389<br>(34.9) | 233 (6.0) | 1 (0.2) | 179 (7.7) | 12 (1.7) | 0 (0.0) | <0.001 |
| 2 | 20544<br>(53.6) | 2319<br>(60.0) | 92 (22.5) | 907<br>(38.9) | 226<br>(32.4) | 42 (8.7) |  |
| 3 | 4142<br>(10.8) | 1218<br>(31.5) | 252<br>(61.6) | 1034<br>(44.4) | 377<br>(54.0) | 309<br>(64.0) |  |
| 4 | 226 (0.6) | 93 (2.4) | 63 (15.4) | 175 (7.5) | 77 (11.0) | 131<br>(27.1) |  |
| 5 | 47 (0.1) | 5 (0.1) | 1 (0.2) | 34 (1.5) | 6 (0.9) | 1 (0.2) |  |
| *QCG scores (median [IQR])* |  |  |  |  |  |  |  |
| ACS | 1.04<br>[0.22,<br>3.28] | 2.80<br>[1.18,<br>6.22] | 5.70<br>[3.00,<br>10.74] | 13.05<br>[2.30,<br>25.76] | 8.30<br>[2.77,<br>23.89] | 16.17<br>[5.20,<br>30.90] | <0.001 |
| STEMI | 0.01<br>[0.00,<br>0.04] | 0.03<br>[0.01,<br>0.14] | 0.13<br>[0.03,<br>0.60] | 0.34<br>[0.05,<br>1.51] | 0.40<br>[0.06,<br>1.65] | 1.13<br>[0.20,<br>5.93] | <0.001 |
| Myocardial injury | 1.32<br>[0.39, | 3.16<br>[1.47, | 5.90<br>[3.33, | 16.82<br>[5.80, | 14.22<br>[6.28, | 18.89<br>[9.78, | <0.001 |
|  | 3.46] | 6.10] | 9.68] | 25.89] | 23.58] | 32.11] |  |
| Pulmonary edema | 0.49<br>[0.16, 1.50] | 1.51<br>[0.53, 4.02] | 4.13<br>[1.37, 9.03] | 12.02<br>[1.72, 25.26] | 18.05<br>[4.34, 26.97] | 22.21<br>[9.31, 36.40] | <0.001 |
| Pericardial effusion | 0.02<br>[0.00, 0.14] | 0.06<br>[0.01, 0.45] | 0.26<br>[0.03, 1.51] | 0.89<br>[0.05, 6.10] | 2.48<br>[0.22, 9.80] | 3.11<br>[0.67, 9.40] | <0.001 |
| Left ventricular dysfunction | 0.06<br>[0.02, 0.21] | 0.15<br>[0.04, 0.61] | 0.95<br>[0.27, 3.80] | 3.07<br>[0.47, 16.05] | 3.50<br>[0.72, 13.14] | 21.00<br>[5.46, 54.70] | <0.001 |
| Right ventricular dysfunction | 0.10<br>[0.03, 0.36] | 0.26<br>[0.08, 1.10] | 1.01<br>[0.30, 3.60] | 3.80<br>[0.48, 15.12] | 5.62<br>[1.11, 16.47] | 11.08<br>[3.90, 26.73] | <0.001 |
| Pulmonary hypertension | 0.11<br>[0.03, 0.48] | 0.40<br>[0.09, 1.72] | 1.40<br>[0.30, 5.32] | 4.05<br>[0.37, 15.08] | 7.90<br>[1.43, 19.79] | 11.00<br>[3.41, 25.68] | <0.001 |

Group 4 comprised patients classified as high-risk by AI-ECG who underwent surgery without preoperative assessment. This group had the highest frequency of emergency procedures (N = 389); otherwise, most cases were categorized as low-risk surgery by the RCRI (N = 1,991) or ESC surgical risk (N = 1,147) (**Figure S4** in Multimedia Appendix). Among the remaining patients (N = 137) after excluding these two reasons, 105 underwent preoperative CV testing at another hospital. Thus, the number of “true” missing cases by clinicians was 32, in which no further diagnostic tests or cardiology consultations were undertaken despite high-risk AI-ECG findings. Among these 32 cases, two deaths occurred (mortality rate: 6.25%).

Group 5 included cases classified as high risk by AI-ECG, but with negative preoperative CV tests. This group consisted of a heterogeneous set of conditions: (1) abnormalities detected by preoperative CV testing that were not classified as positive results (e.g., atrial fibrillation, myocardial disease, and severe left ventricular hypertrophy); (2) a history of open-heart surgery but with preserved cardiac function; or (3) stable coronary stenosis or prior percutaneous coronary intervention without interval change. Group 5 had the highest composite endpoint rate (6.3%).

Group 2 included cases with possibly avoidable tests performed because of the AI-ECG low-risk stratification and negative preoperative CV tests. The most frequently performed test modality in this group was echocardiography (91.9%), which resulted in the highest cost of preoperative CV tests among the six study groups (**Figure 4**). The proportion of the cost spent in Group 2 was 57.8% of the total spending on preoperative CV surgeries.

**Figure 4.**
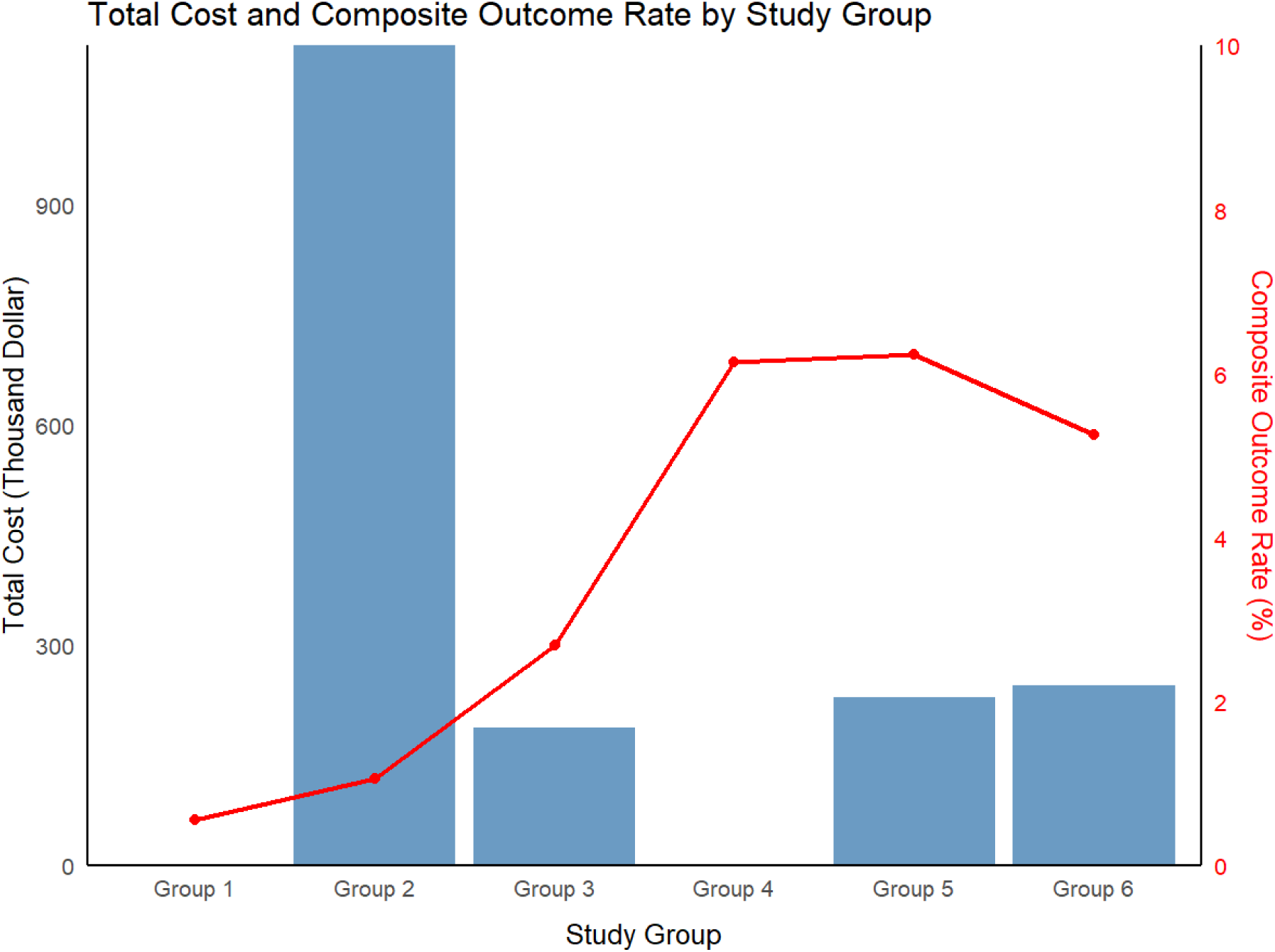
Total cost and composite outcome rate by six study groups. The blue bar graph represents the total cost (thousand USD) and the red line graph represents the composite outcome rate (%).

Group 3 is characterized by the potential “missed cases” when relying solely on AI-ECG. In this group, composite outcomes occurred in 11 patients (2.7%), including one death and 11 cases of prolonged mechanical ventilation. Apart from one craniectomy for intracerebral hematoma evacuation, the remaining 10 procedures were classified as high-risk by the ESC surgical risk category as follows: seven ascending aortic surgeries, one peripheral arterial surgery, one abdominal aortic aneurysm repair, and one pylorus-preserving pancreatoduodenectomy (PPPD). Ten of the eleven surgeries were followed by sequential procedures during the index admission, of which eight were unplanned. A single mortality occurred in a patient undergoing PPPD who had a history of non-ST-elevation myocardial infarction 3 months before surgery.

## Discussion

In this study, we demonstrated the potential utility of multi-task AI-enhanced ECG in reducing potentially avoidable tests and the associated costs. Among patients undergoing non-cardiac surgery, >90% were classified as low risk by AI-ECG. Notably, in those who underwent preoperative CV testing despite a low-risk AI-ECG classification, 90.5% had negative results; however, this negative-test group incurred more than half of the overall expenditure for preoperative CV testing in all surgeries. When the AI-ECG risk classification was integrated with traditional risk-stratification tools, such as the ESC surgical risk category and RCRI, patients identified as low-risk had an extremely low incidence of postoperative composite outcomes. This potentially enables the conservation of medical resources by minimizing potentially avoidable advanced CV evaluation.

The indiscriminate use of preoperative CV tests has raised several important concerns. Overuse not only increases healthcare costs and delays surgery but may also postpone tests intended for diagnosing other urgent conditions. Furthermore, the prevalence of serious cardiac diseases that require a change in surgical plan is low in elective surgery candidates unless they have cardiac symptoms or signs, resulting in a low yield of meaningful results.[23] Nevertheless, preoperative CV tests, especially echocardiography, are still being performed even before low-risk surgery in a substantial number of cases. In a retrospective study of low-risk surgery, 39.8% of patients underwent echocardiography, among which only 5% showed abnormal results.[24] Moreover, among the diverse abnormal findings that could be obtained from echocardiography, the guidelines for perioperative cardiac management need only left and right ventricular function as preoperative CV diagnostic testing. Consequently, echocardiography is not recommended routinely in asymptomatic and clinically stable patients owing to the lack of benefit.[5] In the meanwhile, no single CV test can fully assess perioperative risk. For example, normal echocardiography findings cannot reliably exclude ischemic heart disease without complementary stress tests. Importantly, postoperative cardiac complications are heterogeneous, ranging from ACS requiring unplanned PCI to heart failure resulting in prolonged ICU stay and even mortality, which cannot be predicted by a single diagnostic modality. Taking together, these limitations highlight the unmet need for a novel and more precise risk-stratification tool to identify high-risk surgical candidates who require further preoperative CV evaluation. Such a tool would enable the more appropriate use of limited healthcare resources, reduce potentially avoidable testing, and ultimately improve perioperative outcomes.

In this study, we introduced a multi-task AI-enabled ECG, originally developed to detect patients at risk of a wide spectrum of cardiac conditions, including STEMI, ACS, left and right ventricular dysfunction, pulmonary edema, pulmonary hypertension, and pericardial effusion, as a novel tool to identify surgical candidates at a low risk of composite postoperative outcomes. When all eight QCG scores were below the optimal threshold, we designated this surgery as low risk. To evaluate the clinical utility of this approach, we categorized all surgery–ECG pairs into three groups according to the results of the preoperative CV testing: No-test, Negative-test, and Positive-test. Each group was further stratified according to the AI-ECG risk classification (low-risk vs. high-risk), resulting in six subgroups. Assuming that the decision to perform preoperative cardiovascular testing strictly followed the AI-ECG risk stratification (i.e., tests performed only in high-risk patients), the number of surgeries in which cardiovascular tests were performed (groups 2, 3, 5, and 6; N = 5,458) exceeded the number of surgeries in which testing would have been required (groups 4, 5, and 6; N = 3,510). This discrepancy corresponded to at least a 35.7% reduction in the number of potentially potentially avoidable tests (**Figure 1**). Given that some patients may require multiple CV tests before non-cardiac surgery, preoperative evaluation using multi-task AI-ECG could be more practical by providing multiple disease-specific risk scores simultaneously from a single ECG image. In contrast to other machine learning algorithms that rely on numerous clinical variables from EHR to predict postoperative adverse events,[25–27] our platform enables simple and efficient on-site risk stratification using only a single ECG.

In group-wise analysis, Group 2 emerged as the most notable subgroup. Despite being classified as low-risk by AI-ECG and demonstrating a low prevalence of composite outcomes, this group (N = 3,868) accounted for more than half of the total patient expenses. These patients underwent potentially avoidable preoperative cardiovascular testing, whereas Group 4 (N = 2,329), characterized by a high AI-ECG risk, did not undergo testing despite their higher event rate. Taken together, approximately 40% of the cardiovascular tests were potentially potentially avoidable, and the composite outcome incidence was markedly higher in Group 4 than in Group 2. This finding underscores that AI-ECG not only decreases redundant testing but also provides more appropriate risk stratification.

Group 3 represents another potentially “missed” population under an AI-ECG–guided strategy. Given the retrospective nature of the study, allocating patients for further CV testing was conducted based on the physician’s evaluation, which implies the potential existence of clinical risk for perioperative complications. Namely, the higher adverse event rates in group 3 compared to groups 1 and 2 could be the result of further stratification within the low-risk AI-ECG group based on clinical information. Moreover, most adverse events that developed in this group were neither predictable nor preventable by preoperative testing. On the other hand, in cases without composite outcomes, 39 patients with abnormal coronary angiography underwent revascularization (PCI or CABG) prior to surgery, and 4 patients with severe aortic stenosis underwent surgery with intensive monitoring. **Figure S5** in Multimedia Appendix presents a representative case of Group 3, demonstrating a significant change in AI-ECG scores before and after PCI. Given that the ECG closest to the index surgery was selected to determine the AI-ECG risk group, such interventions may have helped lower perioperative risk and prevent adverse postoperative events.

Group 5 provides meaningful insights. Although this group demonstrated no abnormal findings in preoperative cardiovascular tests, it exhibited the highest rate of composite outcomes among all groups. A large proportion of these patients had undergone prior open-heart surgery, coronary artery bypass grafting, PCI, or stable coronary artery disease. Given the well-recognized elevated perioperative risk in these populations, repeated cardiovascular testing with unchanged results is unlikely to affect intraoperative or postoperative management. Therefore, combined with delicate history-taking about their symptoms, AI-ECG might be able to replace potentially avoidable repeated examinations in clinically stable patients with a history of cardiac surgery or PCI. In addition, conditions overlooked in the preoperative assessment, such as atrial fibrillation or underlying myocardial disease, may have contributed to the adverse outcomes. These factors highlight potential novel targets for reducing postoperative complications beyond the established strategies.

Low-risk patients are potential beneficiaries of AI-ECG risk stratification. Although current guidelines do not recommend performing additional tests in patients undergoing low-risk surgery not otherwise indicated by corresponding symptoms and/or signs of heart disease,[5, 6] a substantial number of patients undergoing low-risk surgeries are preceded by preoperative CV tests in the real world.[24] In our study, patients with low risk (i.e., having RCRI scores of 0 or 1 and undergoing low-risk surgeries by ESC surgical risk category) had very low composite outcome rates with high RR (**Table 2**). Namely, an AI-enabled ECG could potentially reduce unnecessary tests, at least in patients with low surgical risk and low-risk AI-ECG. Therefore, integrating AI-ECG with established risk tools may outline patients who benefit from preoperative CV testing while curtailing waste of medical resources.

Emergency or urgent surgeries could also benefit from an AI-enabled ECG, in which the time for the preoperative workup is usually limited. As the risk of delaying surgery generally exceeds the benefit of information from preoperative examinations, risk assessment has been omitted for time-sensitive surgery.[28] However, the risk of developing composite outcomes in emergency surgeries is much higher than that in elective surgeries. If we could obtain information about the patient’s critical cardiac condition without delay, the results might change with intensive monitoring and delicate postoperative care. In patients who underwent emergency surgery, the risk of the composite endpoint was 6.5% in the low-risk group and 25.8% in the high-risk group on AI-ECG. Therefore, medical resources for postoperative care can be reassigned according to the rapid risk classification by AI-ECG, particularly in resource-limited settings.

From a cost perspective, preoperative risk stratification using AI-ECG may offer significant benefits. In our cohort, 5,458 patients underwent preoperative cardiovascular testing, whereas only 3,510 were classified as high risk by AI-ECG. Thus, approximately 35.7% of patients could have undergone additional testing, with corresponding savings in medical resources and costs. However, these findings should be interpreted in the context of each country’s healthcare system, including the costs of individual tests, waiting times, and insurance coverage. In South Korea, where universal health insurance facilitates easy access to diagnostic testing, physicians may prescribe more tests than those recommended by the guidelines, potentially leading to an overestimation of costs. Furthermore, because downstream costs related to beneficial or adverse events were not included, a prospective cost-effectiveness analysis is necessary. Nevertheless, given its long-standing role in preoperative assessment, the ability of AI-ECG to refine risk stratification and reliably identify low-risk patients without additional expense may represent a meaningful advantage.

This study had some limitations. First, as a single-center retrospective analysis, the proportion of low-risk surgeries was relatively high, and the overall mortality rate was lower than that reported in previous studies. Second, the unique insurance system in South Korea may have influenced the frequency and cost of preoperative testing, limiting the generalizability of cost-related findings. Third, although AI-ECG risk stratification was associated with postoperative outcomes, it remains unclear whether its use, followed by appropriate interventions, would improve clinical outcomes or reduce overall expenses. Because preoperative testing was not randomly assigned and reflects clinical judgment, the observed associations should not be interpreted causally. Fourth, stress testing was not included as a preoperative CV test, which may have led to an underestimation of the scope of conventional testing. However, completing preoperative risk assessment using stress testing demands more time and cost without a significant increase in diagnostic ability.[29] Finally, patient symptoms, signs, and functional capacity were not incorporated, and in patients with suspected but undiagnosed cardiac disease, further evaluation remains necessary regardless of AI-ECG classification. To overcome these limitations, prospective randomized controlled trials should be conducted for the comprehensive assessment of whether AI-ECG can improve postoperative outcomes and reduce the cost related to preoperative CV tests in non-cardiac surgery.

In conclusion, using multi-task AI-ECG is associated with reducing potentially avoidable preoperative cardiovascular testing and the associated costs without increasing the incidence of postoperative adverse events. By integrating AI-ECG with established risk-stratification tools, a new low-risk group with an extremely low event rate can be identified, for whom additional testing may not be required. These findings suggest that AI-ECG may serve as a cost-effective adjunct to current preoperative assessment strategies in non-cardiac surgery.

## Acknowledgment

None

## Data Availability

The datasets analyzed during the current study are not publicly available due to institutional restrictions but are available from the corresponding author on reasonable request.

## Author Contributions

All authors were involved in the conceptualization and design of this research. YK, JP, ICH, and GYC collected the data, and HMC, YK, JK, JHL, and IYO analyzed the data. JP, ICH, YEY, YYC, and JHL conducted the investigation. JK and YC were responsible for generating the AI-ECG scores. HMC and YK drafted the manuscript and prepared the figures; YEY, JK, YC, and IAS reviewed and revised the manuscript. All the authors approved the final manuscript.

## Abbreviations

ACS: Acute coronary syndrome
AI: artificial intelligence
AUROC: area under the receiver operating characteristic curve
CAG: coronary angiography
CCTA: coronary computed tomography angiography
CI: confidence interval
CV: cardiovascular
ECG: Electrocardiography
EHR: electrical healthcare records
ESC: European Cardiology of Society
PCI: percutaneous coronary intervention
PPPD: pylorus-preserving pancreatoduodenectomy
QCG: quantitative score for ECG
RCRI: Revised Cardiac Risk Index
RD: risk difference
RR: relative risk
SPECT: single-photon emission computed tomography
STEMI: ST-segment elevation myocardial infarction

## Multimedia Appendix

**Table S1. The list of QCG scores used to define the risk by AI-enabled ECG**

**Table S2. Characteristics according to the preoperative cardiovascular test groups**

**Table S3. Comparison of preoperative cardiovascular (CV) tests and the results stratified by study groups**

**Figure S1. Study flow**

**Figure S2. The distribution of QCG scores.** (A) Histograms for the distribution of individual QCG scores. Red dashed line represents the optimal cutoffs for individual scores. (B) The distribution of QCG scores across the six study groups. Low-risk AI-ECG group was illustrated in light blue, while high-risk AI-ECG group with orange.

**Figure S3. Composite outcome rates according to the reclassification using AI-ECG.** (A) The European Society of Cardiology (ESC) surgical risk categories and (B) the Revised Cardiac Risk Index (RCRI) are reclassified by AI-ECG.

**Figure S4. The explainable causes of surgery without further preoperative cardiovascular testing in group 4.**

**Figure S5. The representative case with abnormal coronary angiography (CAG) results requiring revascularization before proceeding to surgery.** (A) ECG and AI-ECG results before CAG; (B) the closest ones before surgery; (C) CAG findings; (D) CAG after PCI.

