## Supplementary file for "Multitask Artificial Intelligence–Based Electrocardiogram Tool for Preoperative Cardiac Testing in Noncardiac Surgery: Retrospective Cohort Study of Health Care Utilization and Costs"

**Table S1. The list of QCG scores used to define the risk by AI-enabled ECG**

| QCG scores | Definition | Youden threshold | Sensitivity | Specificity |
|---|---|---|---|---|
| ACS | Risk of acute coronary syndrome (STEMI, NSTEMI, and UA) | 19.7 | 76.9% | 94.2% |
| STEMI | Risk of ST-segment elevation myocardial infarction developed within 12 hours | 6.6 | 97.5% | 99.0% |
| Myocardial injury | Risk of elevated cardiac troponin | 19.1 | 76.2% | 89.7% |
| Pulmonary edema | Risk of pulmonary edema | 18.1 | 86.0% | 86.5% |
| Pericardial effusion | Risk of large amount of pericardial effusion | 24.6 | 85.3% | 91.0% |
| LV dysfunction | Risk of LVEF < 40% | 30.1 | 90.4% | 87.8% |
| RV dysfunction | Risk of RV dysfunction | 26.0 | 91.6% | 81.5% |
| Pulmonary hypertension | Risk of moderate or severe pulmonary hypertension | 29.3 | 83.0% | 79.1% |

**Table S2. Characteristics according to the preoperative cardiovascular test groups**

| | No-test (N=40,677) | Negative-test (N=4,566) | Positive-test (N=892) | p |
|---|---|---|---|---|
| Demographics and comorbidities | | | | |
| Age (mean (SD)) | 54.80 (15.83) | 69.24 (11.92) | 71.71 (11.23) | <0.001 |
| Male sex (%) | 17745 (43.6) | 2219 (48.6) | 597 (66.9) | <0.001 |
| DM (%) | 4630 (11.4) | 968 (21.2) | 306 (34.3) | <0.001 |
| DM treated with insulin (%) | 411 (1.0) | 159 (3.5) | 62 (7.0) | <0.001 |
| Hypertension (%) | 8492 (20.9) | 1829 (40.1) | 438 (49.1) | <0.001 |
| Previous heart failure (%) | 110 (0.3) | 49 (1.1) | 251 (28.1) | <0.001 |
| Previous ischemic heart disease (%) | 990 (2.4) | 345 (7.6) | 437 (49.0) | <0.001 |
| Previous cerebrovascular accident (%) | 1164 (2.9) | 260 (5.7) | 101 (11.3) | <0.001 |
| Smoking (%) | | | | |
| Nonsmoker | 24730 (60.8) | 3217 (70.5) | 489 (54.8) | <0.001 |
| Smoker | 8299 (20.4) | 1234 (27.0) | 371 (41.6) | |
| Unknown | 7648 (18.8) | 115 (2.5) | 32 (3.6) | |
| Creatinine (median [IQR]) | 0.89 (0.96) | 1.07 (1.25) | 1.65 (1.95) | <0.001 |
| Creatinine ≥ 2.0 mg/dl (%) | 1028 (2.5) | 243 (5.3) | 143 (16.0) | <0.001 |
| Surgery-related | | | | |
| Emergency surgery (%) | 2619 (6.4) | 132 (2.9) | 55 (6.2) | <0.001 |
| Anesthesia (%) | | | | |
| General anesthesia | 29263 (71.9) | 3490 (76.4) | 633 (71.0) | <0.001 |
| Spinal/Epidural anesthesia | 3387 (8.3) | 792 (17.3) | 93 (10.4) | |
| Monitored anesthesia care | 8027 (19.7) | 284 (6.2) | 166 (18.6) | |
| ESC surgical risk category (%) | | | | |
| Low | 23245 (57.1) | 1286 (28.2) | 294 (33.0) | <0.001 |
| Intermediate | 13561 (33.3) | 2513 (55.0) | 318 (35.7) | |
| High | 3871 (9.5) | 767 (16.8) | 280 (31.4) | |
| RCRI (%) | | | | |
| 0 | 26512 (65.2) | 2111 (46.2) | 115 (12.9) | <0.001 |
| 1 | 13127 (32.3) | 2005 (43.9) | 336 (37.7) | |
| 2 | 885 (2.2) | 384 (8.4) | 293 (32.8) | |
| 3 | 123 (0.3) | 59 (1.3) | 126 (14.1) | |
| 4 | 25 (0.1) | 7 (0.2) | 18 (2.0) | |
| 5 | 5 (0.0) | 0 (0.0) | 4 (0.4) | |
| ASA classification (%) | | | | |
| 1 | 13568 (33.4) | 245 (5.4) | 1 (0.1) | <0.001 |
| 2 | 21451 (52.7) | 2545 (55.7) | 134 (15.0) | |

| | | | | |
|---|---|---|---|---|
| 3 | 5176 (12.7) | 1595 (34.9) | 561 (62.9) | |
| 4 | 401 (1.0) | 170 (3.7) | 194 (21.7) | |
| 5 | 81 (0.2) | 11 (0.2) | 2 (0.2) | |
| QCG scores (median [IQR]) | | | | |
| ACS | 3.33 (6.28) | 6.01 (8.26) | 14.93 (17.49) | <0.001 |
| STEMI | 0.21 (1.65) | 0.52 (2.35) | 3.38 (9.22) | <0.001 |
| Myocardial injury | 3.50 (6.27) | 6.37 (8.40) | 16.61 (17.87) | <0.001 |
| Pulmonary edema | 2.29 (5.75) | 5.41 (9.08) | 16.64 (18.32) | <0.001 |
| Pericardial effusion | 0.78 (3.77) | 2.01 (6.62) | 5.10 (9.75) | <0.001 |
| Left ventricular dysfunction | 1.16 (6.08) | 2.46 (7.87) | 19.13 (27.20) | <0.001 |
| Right ventricular dysfunction | 1.23 (4.86) | 2.98 (7.92) | 11.47 (16.88) | <0.001 |
| Pulmonary hypertension | 1.33 (4.56) | 3.55 (7.73) | 11.06 (15.05) | <0.001 |

**Table S3. Comparison of preoperative cardiovascular (CV) tests and the results stratified by study groups**

| | Group 2 (N=3868) | Group 3 (N=409) | Group 5 (N=698) | Group 6 (N=483) | p |
|---|---|---|---|---|---|
| Any preoperative CV test performed (%) | 3868 (100.0) | 409 (100.0) | 698 (100.0) | 483 (100.0) | NA |
| Positive any preoperative CV test (%) | 0 (0.0) | 409 (100.0) | 0 (0.0) | 483 (100.0) | <0.001 |
| Echocardiography performed (%) | 3553 (91.9) | 293 (71.6) | 601 (86.1) | 361 (74.7) | <0.001 |
| Positive echocardiography results (%) | 0 (0.0) | 246 (84.0) | 0 (0.0) | 328 (90.9) | <0.001 |
| Moderate or severe valvular heart disease (%) | 0 (0.0) | 114 (38.9) | 0 (0.0) | 121 (34.1) | <0.001 |
| Regional wall motion abnormality (%) | 0 (0.0) | 133 (45.4) | 0 (0.0) | 241 (68.1) | <0.001 |
| Heart failure (%) | 0 (0.0) | 55 (18.8) | 0 (0.0) | 170 (47.4) | <0.001 |
| LV ejection fraction (%) | 63.33 [60.00, 66.67] | 59.60 [52.33, 65.17] | 61.86 [58.46, 66.00] | 51.01 [41.51, 59.63] | <0.001 |
| Coronary angiography performed (%) | 77 (2.0) | 132 (32.3) | 54 (7.7) | 207 (42.9) | <0.001 |
| Positive coronary angiography results (%) | 0 (0.0) | 102 (77.3) | 0 (0.0) | 171 (82.6) | <0.001 |
| Coronary CT angiography performed (%) | 284 (7.3) | 130 (31.8) | 68 (9.7) | 97 (20.1) | <0.001 |
| Positive coronary CT angiography results (%) | 0 (0.0) | 102 (80.3) | 0 (0.0) | 63 (67.7) | <0.001 |
| SPECT performed (%) | 128 (3.3) | 25 (6.1) | 51 (7.3) | 41 (8.5) | <0.001 |
| Positive SPECT results (%) | 0 (0.0) | 3 (12.0) | 0 (0.0) | 4 (9.8) | <0.001 |

**Figure S1. Study flow**

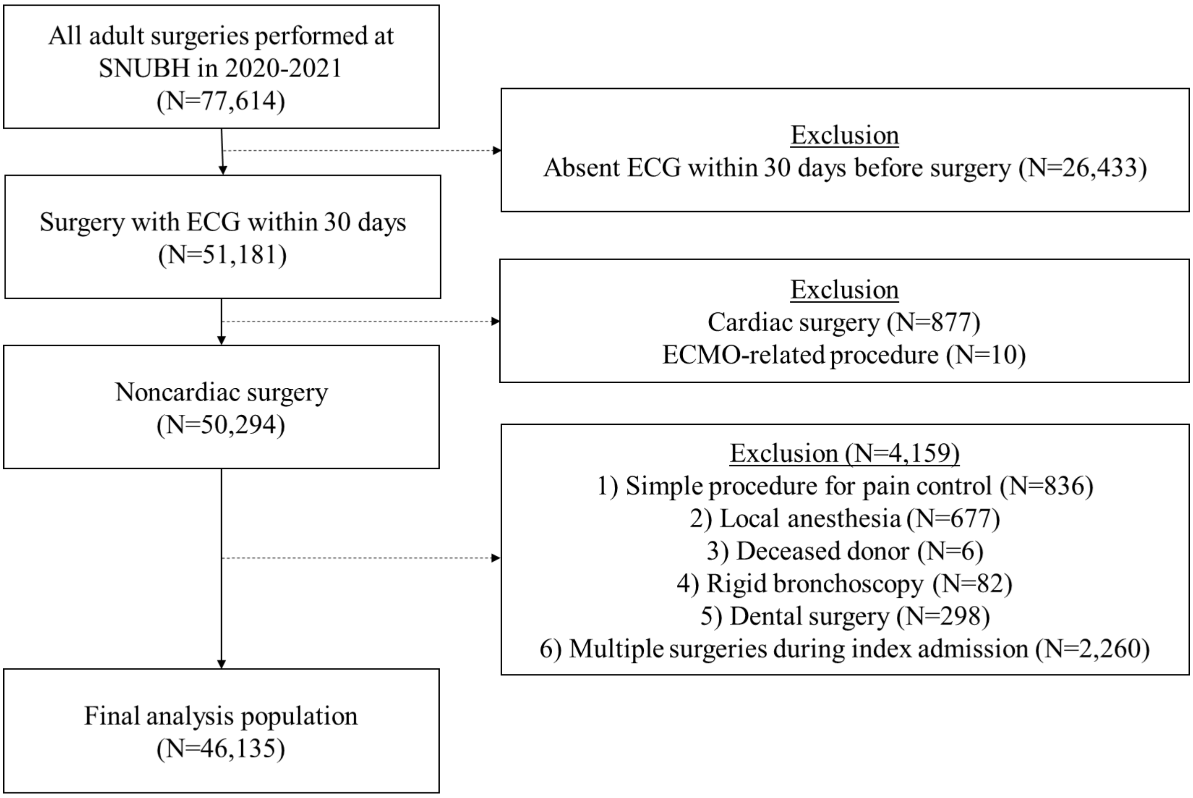

**Figure S2. The distribution of QCG scores.** (A) Histograms for the distribution of individual QCG scores. Red dashed line represents the optimal cutoffs for individual scores. (B) The distribution of QCG scores across the six study groups. Low-risk AI-ECG group was illustrated in light blue, while high-risk AI-ECG group with orange.

**(A)**

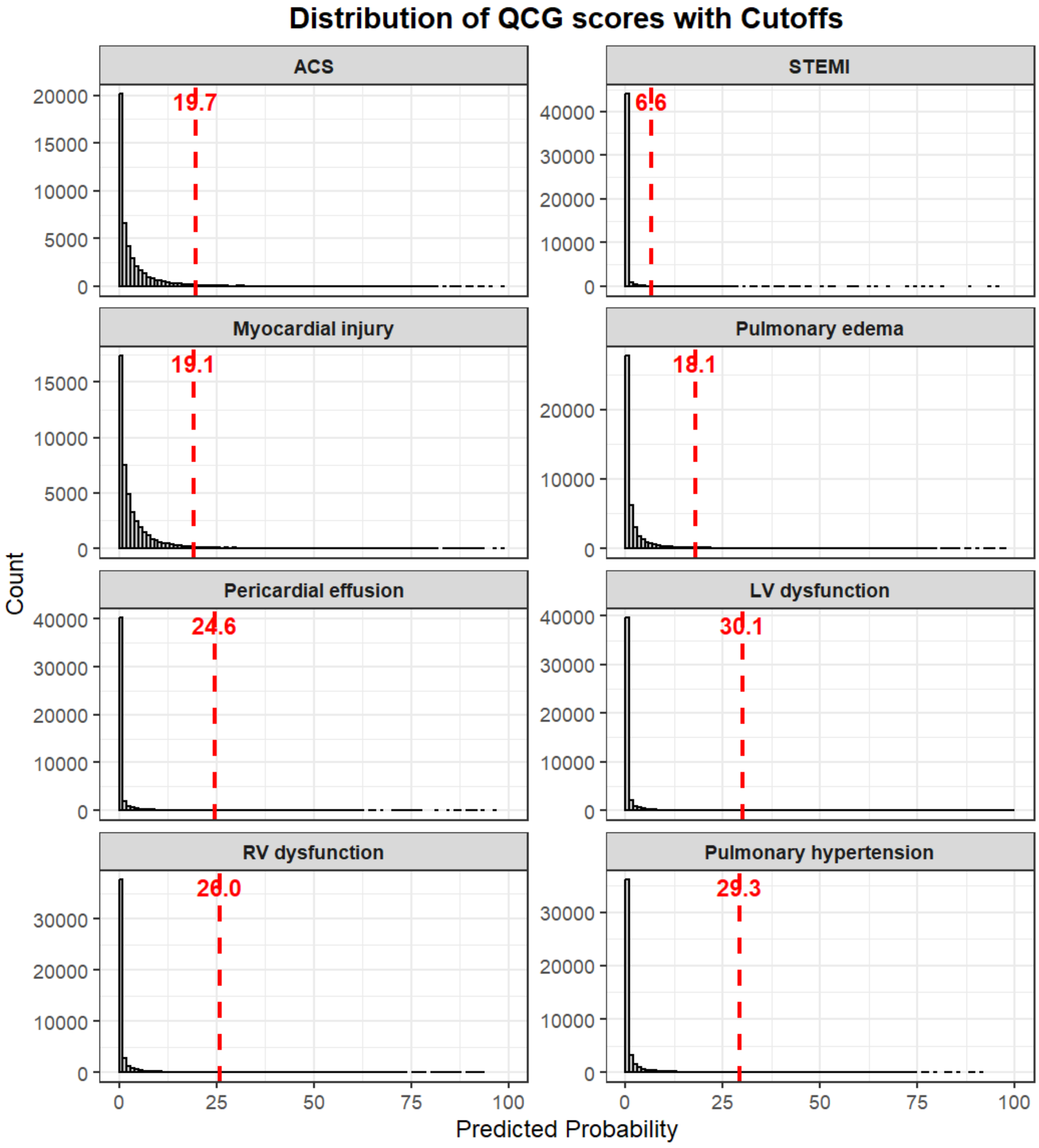

(B)

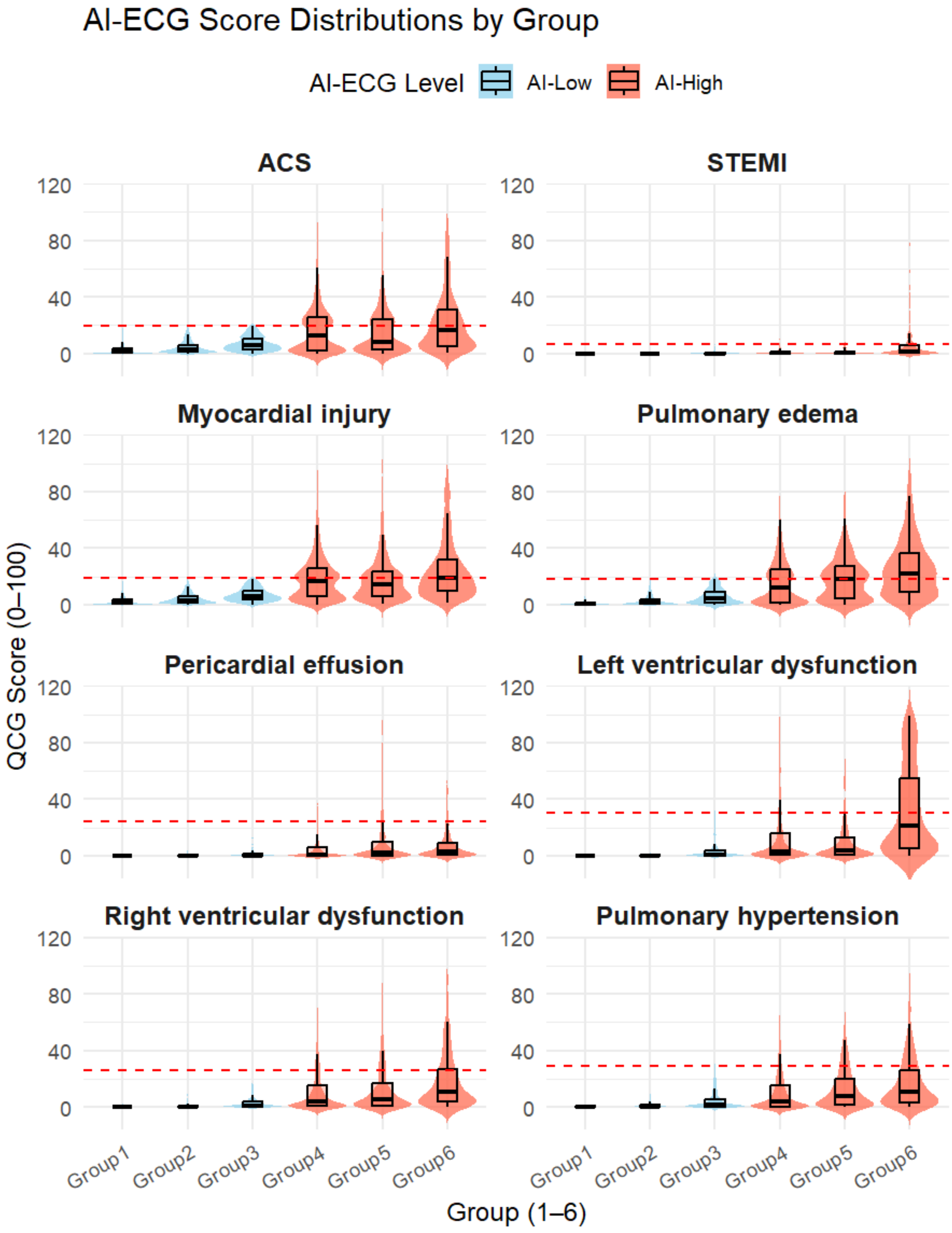
AI-ECG Score Distributions by Group
AI-ECG Level
AI-Low
AI-High
ACS
STEMI
Myocardial injury
Pulmonary edema
Pericardial effusion
Left ventricular dysfunction
Right ventricular dysfunction
Pulmonary hypertension
QCG Score (0–100)
Group (1–6)
Group1
Group2
Group3
Group4
Group5
Group6
0
40
80
120

**Figure S3. Composite outcome rates according to the reclassification using AI-ECG.** (A) The European Society of Cardiology (ESC) surgical risk categories and (B) the Revised Cardiac Risk Index (RCRI) are reclassified by AI-ECG.

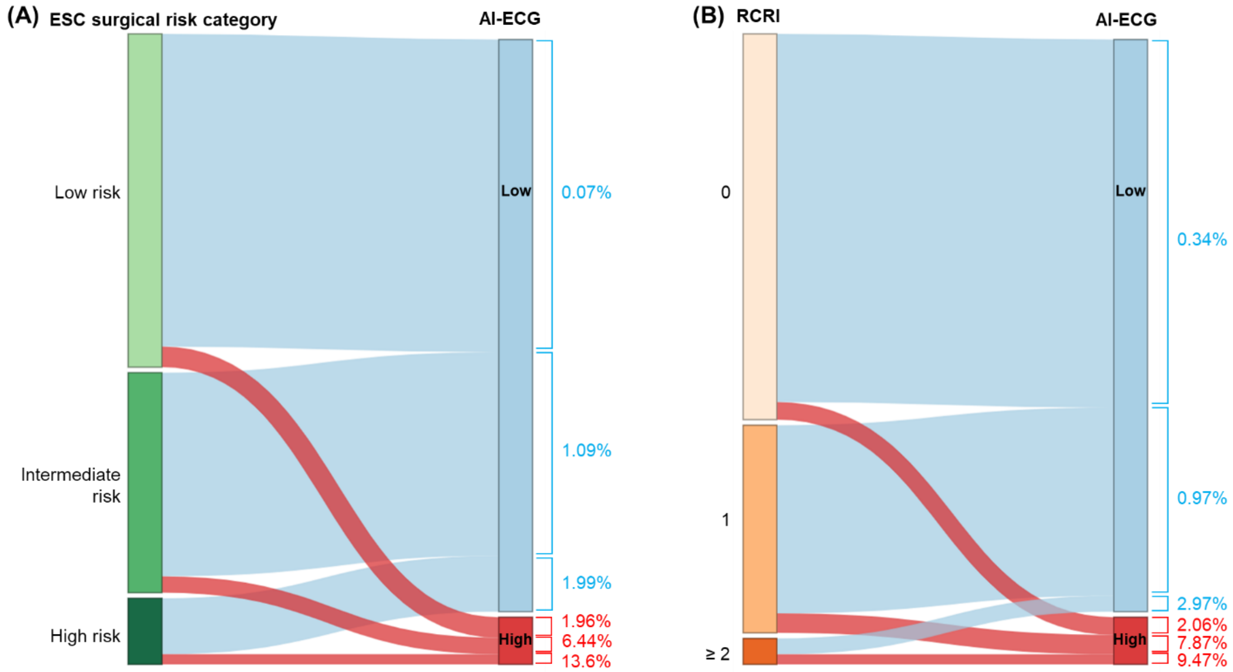

**Figure S4. The explainable causes of surgery without further preoperative cardiovascular testing in group 4.**

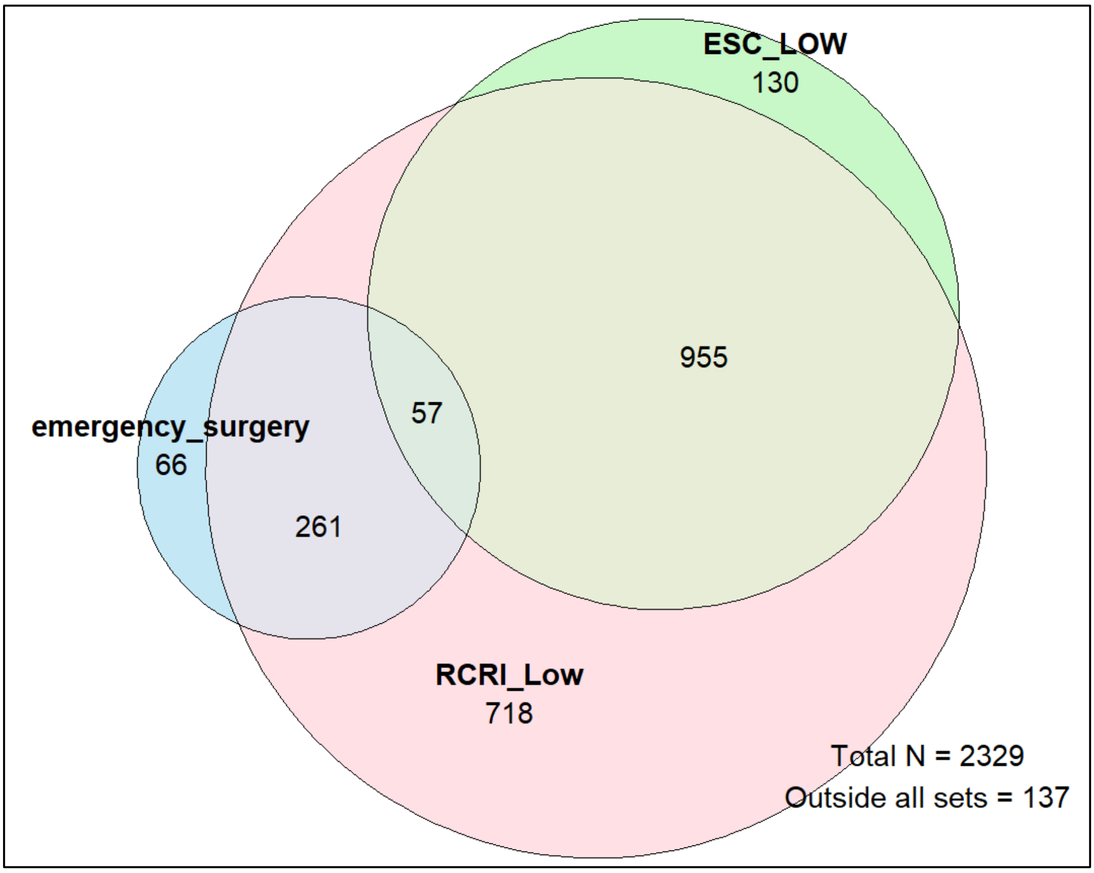

**Figure S5. The representative case with abnormal coronary angiography (CAG) results requiring revascularization before proceeding to surgery.** (A) ECG and AI-ECG results before CAG; (B) the closest ones before surgery; (C) CAG findings; (D) CAG after PCI.

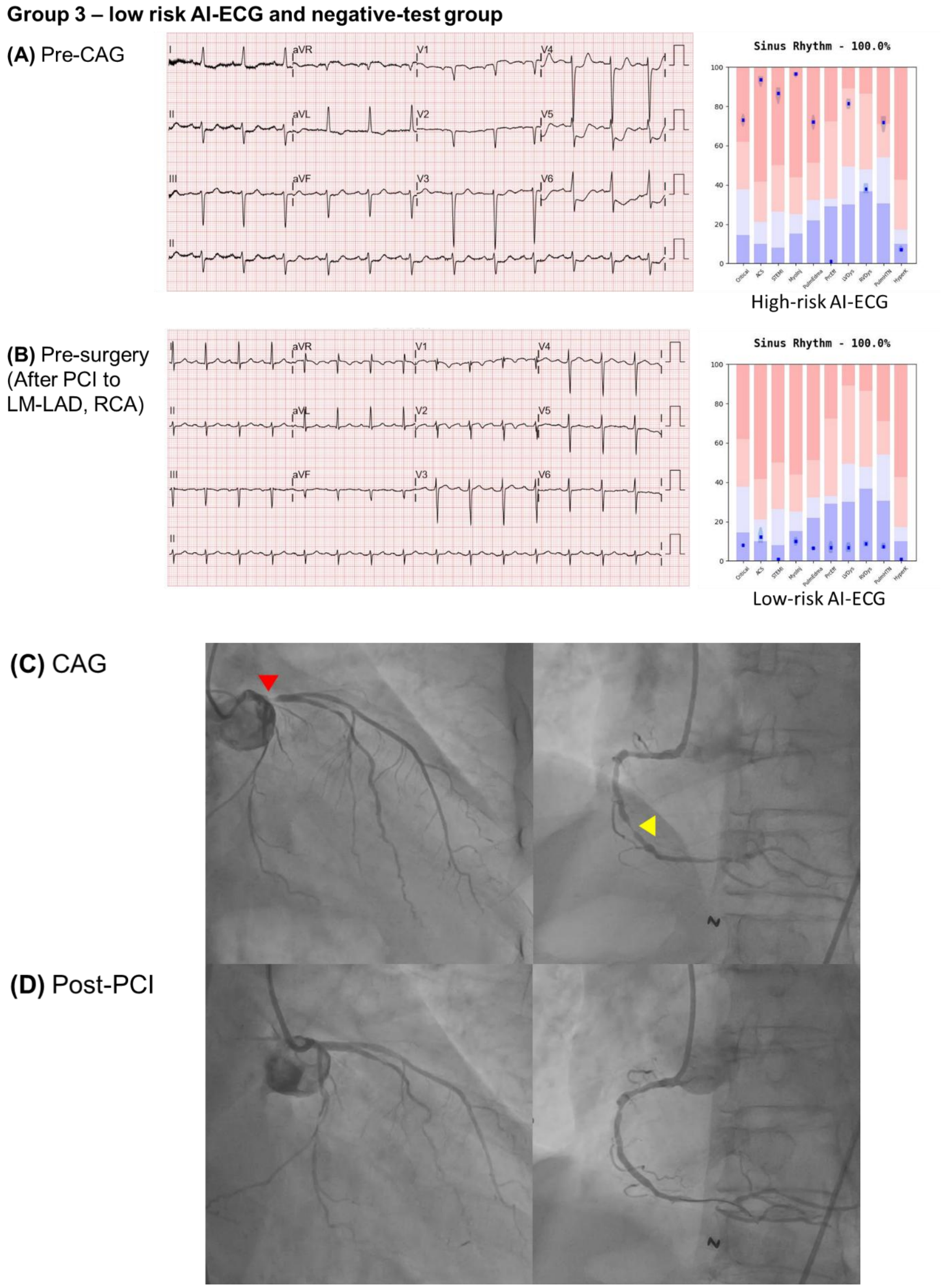
